# A Method For Mimicking Tumour Tissue In Brain Ex-Vivo Ultrasound For Research Application And Clinical Training

**DOI:** 10.1101/2024.09.30.24314649

**Authors:** Alistair Weld, Luke Dixon, Giulio Anichini, Giovanni Faoro, Arianna Menciassi, Sophie Camp, Stamatia Giannarou

## Abstract

**Background:** Intraoperative ultrasound is becoming a common tool in neurosurgery. However, effective simulation methods are limited. Current, commercial, and homemade phantoms lack replication of anatomical correctness and texture complexity of brain and tumour tissue in ultrasound images.

**Methods:** We utilise ex-vivo brain tissue, as opposed to synthetic materials, to achieve realistic echogenic complexity and anatomical correctness. Agar, at 10-20% concentrate, is injected into brain tissue to simulate the tumour mass. A commercially available phantom was purchased for benchmarking.

**Results:** Qualitative analysis is performed by experienced professionals, measuring the impact of the addition of agar and comparing it to the commercial phantom. Overall, the use of ex vivo tissue was deemed more accurate and representative, compared to the synthetic materials-based phantom, as it provided good visualisation of real brain anatomy and good contrast within tissue. The agar tumour correctly produced a region of higher echogenicity with slight diffusion along the margin and expected interaction with the neighbouring anatomy.

**Conclusion:** The proposed method for creating tumour-mimicking tissue in brain tissue is inexpensive, accurate, and simple. Beneficial for both the trainee clinician and the researcher. A total of 576 annotated images are made publicly available upon request.

## Introduction

Intraoperative ultrasound (IOUS) is an increasingly popular tool, as it is a viable alternative to more complex, time consuming and expensive solutions such as intraoperative magnetic resonance imaging [1] [2] However, who performs this task is not well defined, whether it should be the surgeon, radiologist, sonographer, or a combination of these professionals. The background commensurate training, in turn, remains highly variable and not standardised. In radiology, radiologists and sonographers undergo formal training and routinely perform hundreds of supervised ultrasound (US) scans on real patients over a condensed period to reach competency with accepted forms of assessment and accreditation. In contrast, in surgery, US training is highly variable depending on local practice and expertise and typically lacks the same structure. This is compounded by the much lower frequency of cases and greater risks that are inherent to intraoperative US which greatly limits access and slows experience in contrast to routine nonoperative diagnostic US which is high volume, low risk, and ubiquitous across most hospitals. Considering these challenges but also the many strengths of IOUS, such as real-time imaging and ease of integration into the surgical workflow, methods to improve surgical training in IOUS are needed.

A particularly noteworthy application of IOUS is in oncological brain surgery, where the goal is maximal safe resection (removing the most tumour while preserving function and avoiding disability). This is due to the reduced reliability of preoperative imaging and challenges with delineating pathological from functional tissue [3]. Precision and safety are of the utmost importance, as the probe interacts with highly sensitive, fragile, and functionally crucial cortical surfaces. During tumour resection, a craniotomy is performed to expose a localised section of the brain surface directly above the tumour. IOUS can then be used to locate pathological tissue. This cavity is typically small, with a limited field of view, and will often contain blood products, gas, and other sources of artefacts that can complicate scanning [4]. The difficulty with gaining necessary IOUS experience is particularly daunting in neurosurgery, where it is uniquely not possible to perform practice US scanning of the brain on people outside of the operating theatre as a window through the skull is needed. This further steepens the learning curve impacting adoption and performance.

Traditionally, medical phantoms are used to simulate anatomy and medical procedures. There is limited access to purchasable educational tools and known homemade methods for creating phantoms for brain IOUS. Most commercially available brain phantoms are primarily designed for basic surgical training and tend to be expensive. As ultrasound is only one facet of the surgical process, the anatomical correctness of the tissue is usually simplistic, lacking real structural and textural details. This, in the case of the brain, is a significant issue due to the inherent complexity of the anatomy.

Homemade phantoms can be a more affordable and customisable solution. These phantoms can be made using food products [5] such as spam [6] or solutions such as ballistic gelatin [7] [8], bovine gelatin [9], sodium alginate [10], polyvinyl alcohol (PVA) [11] or agar [12] [13]. In particular, agar has been shown to successfully replicate the mechanical properties of human tumour tissue [14]. Although these methods are inexpensive, the core structure of the phantom will be simplistic and homogenous. Foreign bodies can be embedded in the medium to create diversity in echogenic features [15]. However, in combination this isn’t realistic. Real tissue is complex, and the interpretation of the ultrasound perspective and visible anatomy is one of the fundamental challenges in IOUS.

This article highlights the importance of creating anatomical accuracy in phantoms. A method is proposed to create a phantom to mimic tumours in inexpensive brain ex vivo using agar.

## Materials and Methods

A summary of the cost and difficulty of creating the phantom is provided in Table.1. As a benchmark for commercially available phantoms, the ‘Tumour box’ and 5 ‘Glioma Cartridge’ from UpSurgeOn [27], were purchased at the cost of 654.92 € and 855.00 € with a shipping fee of 50.00 €. This phantom is designed primarily for resection practice, with ultrasound property being only a secondary feature. However, this was the reason for the preference of this phantom, as with most other medical training phantoms, the phantom will be interacted with in such a way that there will be destruction of the material. For example, phantoms are commonly used for vascular access, epidurals, joint injections, or aspiration. Therefore, having multiple cartridges is a significant benefit of this phantom, unlike alternatives such as the Adult Brain phantoms by True Phantom Solutions. Although the latter may have better imaging feedback, it costs more ($1,738 Basic, $3,090 Standard and $4,250 Complex model), it is non-replaceable and can only be used in such a way as to preserve the integrity of the material. No cutting, modification, or rough handling.

**Table 1.** Cost, acquisition and setup.

|  | UpSurgeon | Animal Brain |
| --- | --- | --- |
| Cost | High | Low |
| Availability | Moderate | Easy |
| Time to construct | 5 min | 10 min |
| Special storage | No | Yes |
| Special handling | No | Yes |
| Complexity of set up | Simple | Intermediate |

Multiple ovine and bovine ex vivo samples (Figure.1) were purchased at the cost of £5 each. All data were captured using an Ultrasonix Sonix CEP ultrasound system and a L14-5 linear probe. This was chosen as it is a previous generation machine, therefore producing lower quality images than can be expected with the modern state-of-the-art systems and mimicking a real-life scenario where clinicians rarely have access to the latest technological developments. Among the different materials used in the literature, to simulate brain tumours, agar was chosen due to its low cost and matching mechanical properties [14]. Agar is also easy to manipulate, since there is no need for freeze-thawing cycles (as needed with PVA [11]), and it can be injected during its liquid stage and allowed to solidify after injection in minimal time.

**Figure 1.**
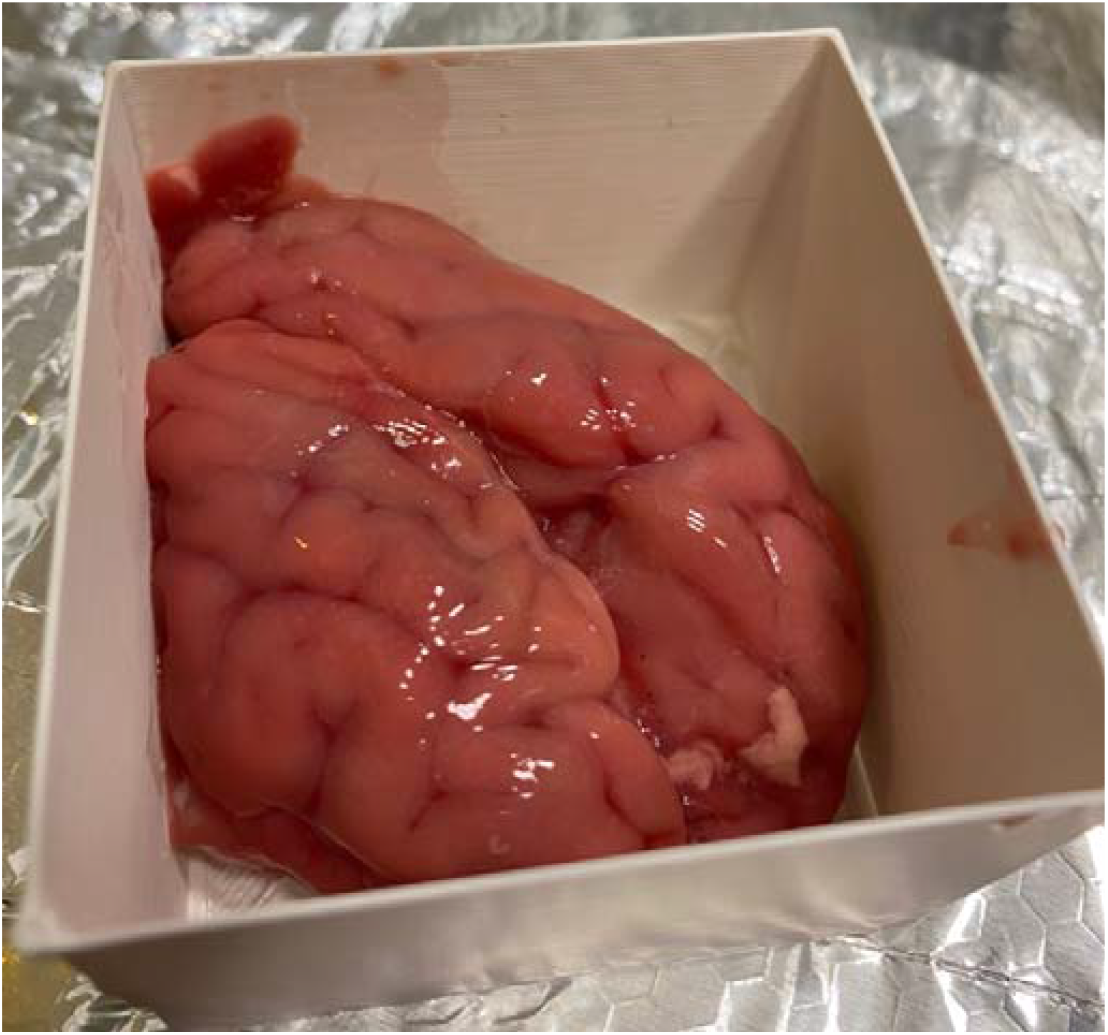
An ovine sample used to collect the data, before the agar solution was injected. The container has the dimensions 6.5cm x 7.0cm.

Tumour simulation was achieved using 2ml of 10-20% concentrate agar, heated using a Fisherbrand™ Hotplate Stirrer set to 150° and 1500 rpm, injected into the depth of the subcortical white matter/centrum semiovale - the agar was cooled before injection. Where both these areas are approximately equivalent to the human counterpart in terms of anatomical features such as texture, locations, colour, and physiological role. Depending on the concentration, temperature, volume, and area injected, from our observations, the time it took to harden could vary anywhere from a few minutes up to half an hour. To simulate the necrotic core of a tumour, water was injected into the hardened agar. Ultrasound feedback was used to guide the needle into the agar solution.

Two experienced IOUS operators individually assessed the phantoms in relation to the acquired images. Evaluation categories were defined for qualitative assessment, using the five-point Liekert scale. Five categories were chosen, which were considered comprehensive for the evaluation of medical phantoms and their similarity to real brain tissue. The categories are Reusability, Echotexture, Cerebral structure, Tactile feedback, and Tumour mimic semblance.

## Results

The results of the qualitative ratings are shown in Table.2. Overall, the quality of the phantoms made using real brain tissue produced superior imaging, anatomical accuracy, and tumour accuracy, in comparison to the phantom. However, the animal phantom was not reusable and had to be disposed of on the day of use to prevent decay and necrosis.

**Table 2.** Qualitative assessment using a five-point Liekert scale (1 = poor to 5 = excellent)

|  | UpSurgeon | Animal Brain |
| --- | --- | --- |
| Reusability | 4 | 2 |
| Echotexture | 3 | 5 |
| Cerebral structure | 1 | 5 |
| Tactile feedback | 2 | 5 |
| Tumour mimic semblance | 2 | 4 |

### Phantom analysis

Figure.2 shows an image of the phantom captured by our device. The most prominent and noticeable features are the linear, horizontal hyperechogenic lines. Although useful in defining the echogenic nature of the tissue analysed, their clinical correlation is debatable. The tumour itself appears as a circular hyperechoic ring with a hypoechoid core, causing significant acoustic shadowing. In the clinical setting, such shadowing is generally indicative of a tumour with a calcified component. In general, although there is echogenic diversity, these features are incoherent and do not resemble real anatomy.

**Figure 2.**
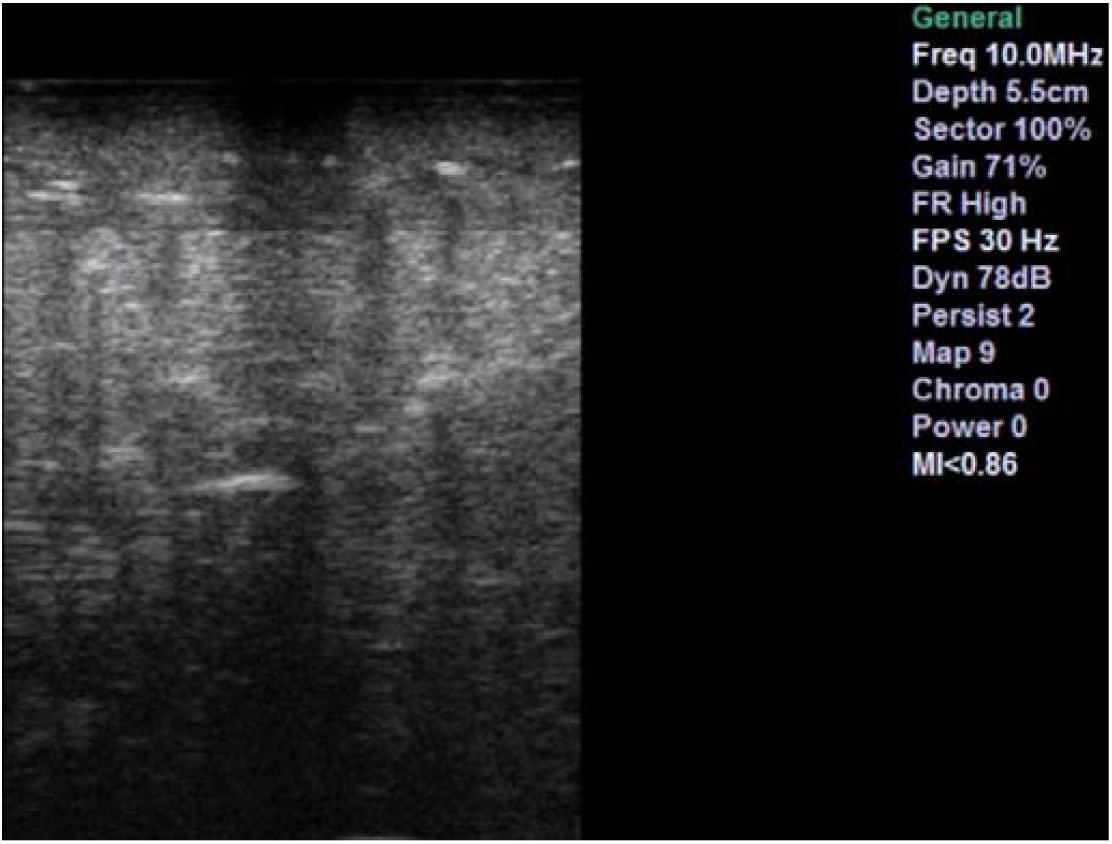
UpSurgeOn brain box phantom image from our machine.

### Ex-Vivo and agar

The unmodified bovine brain sample is shown on the left side of Figure.3 and the ovine sample in section A and B of Figure.4. The good contrast within the tissue is the first noticeable property. The samples provided a good visualisation of the cortical sulci and gyri, with evident arachnoidal hyperechogenity and differentiation of grey-white matter. There were no shadows, artefacts, or elements of degradation. The images obtained through these specimens closely resembled a mammalian brain anatomy and therefore were considered more reliable to reproduce a clinical neurosurgical scenario.

**Figure 3.**
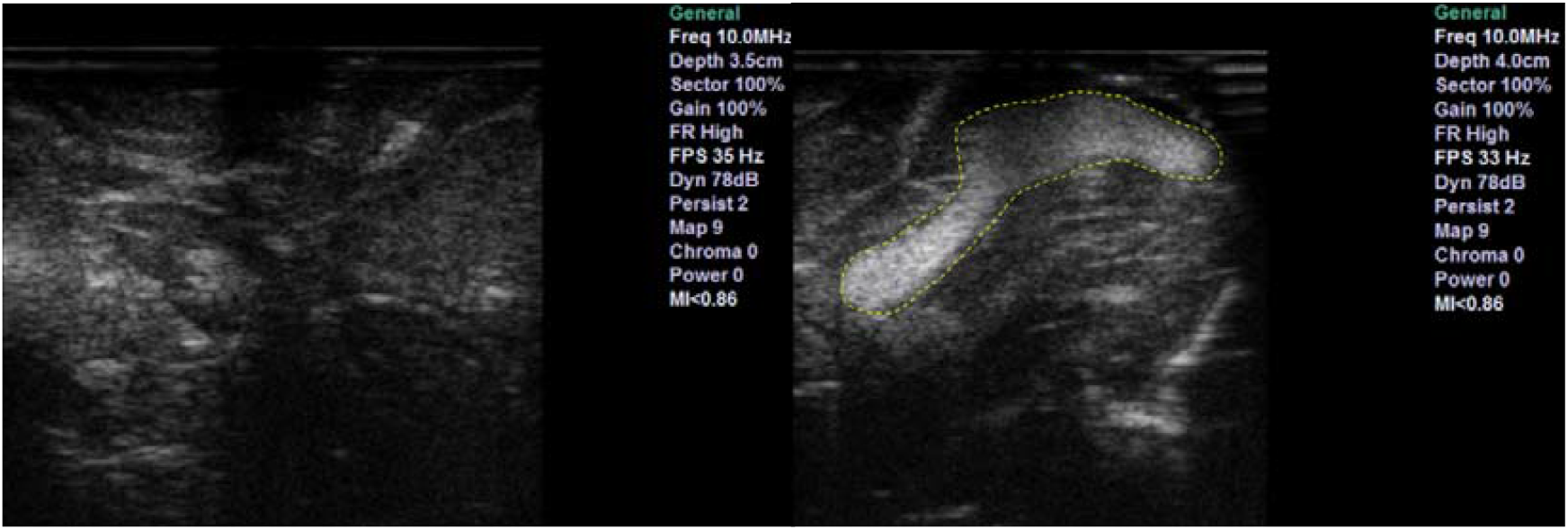
Bovine sample. Left shows ultrasound feedback of phantom without tumour addition. Right shows ultrasound feedback with agar injected

**Figure 4.**
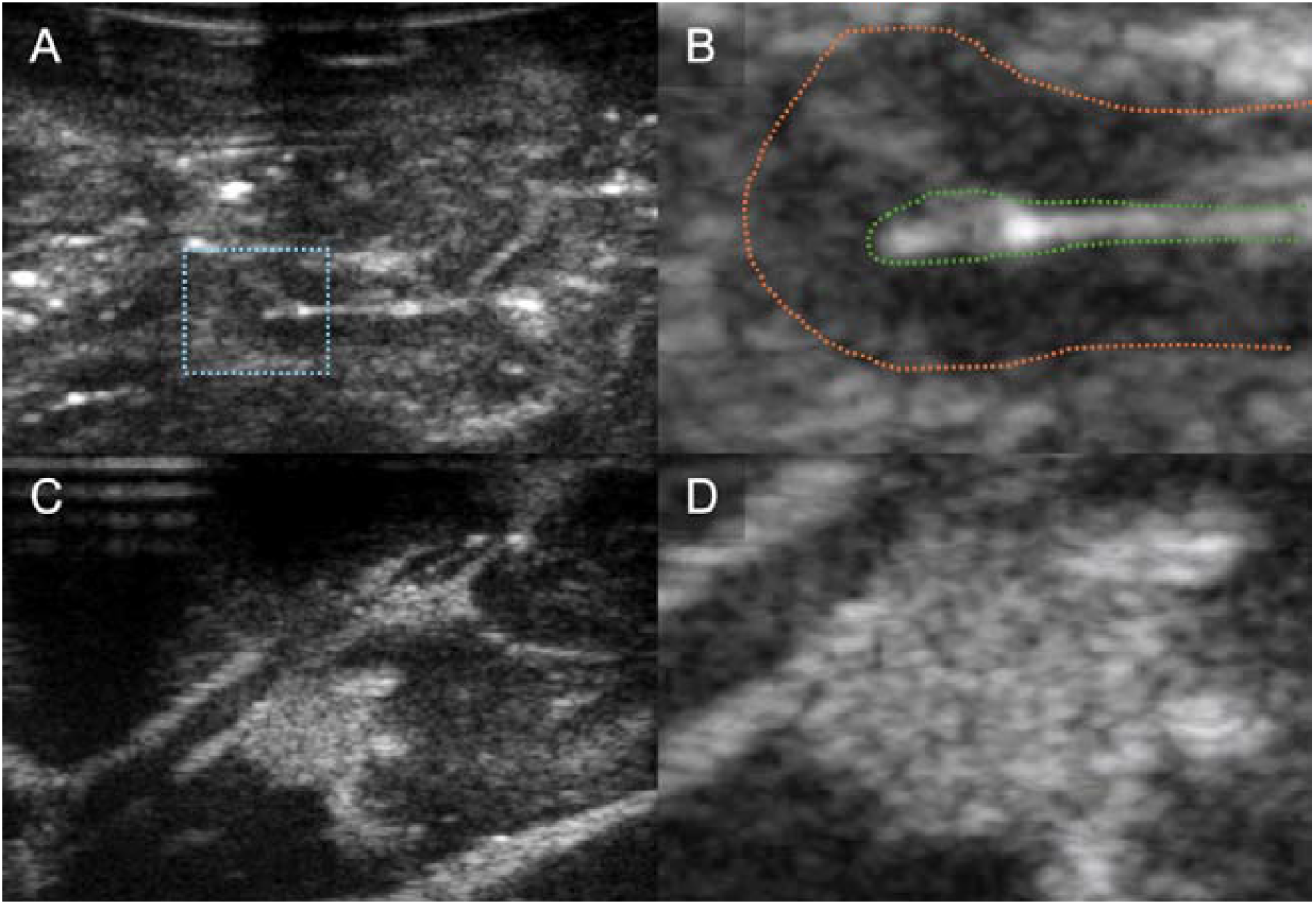
Ovine sample. B-mode image of sheep brain (A) with magnified image of gyrus (B) demonstrating discernible normal cerebral architecture with sulci and pia (outlined by green dotted line), cortex (outlined by orange dotted line) and surrounding white matter. B-mode image of the same sheep brain after localised infiltration of agar to mimic a hypercellular, echogenic tumour (C) with magnified image of tumour mimic demonstrating clear margins (D).

In the same figures, the same tissues are also shown, but with the addition of injected 10-20% concentrate agar. In the context of the bovine brain, the addition of agar is shown in the right image of Figure.3. With the ovine tissue, sections C and D in Figure.4 show the agar solution. The agar solution was injected into the depth of the inferior frontal sulcus. As expected, the tumour appeared embedded in the subcortical white matter creating an echogenic sphere, with a narrow tract following the entry point.

Unlike phantoms, agar injection into a real brain not only showed a well-defined and clearly identifiable anomaly but also provided a mass and an infiltrative effect to the surrounding areas. This interaction with brain tissue appears to be more reliable and useful for educational purposes and research applications.

For reference of a normal IOUS view of brain tumour tissue, Figure.5 shows images of glioblastomas - highlighted with yellow dotted lines - taken intraoperatively and right before resection, at Charing Cross Hospital, using a state-of-the-art system Canon i900 ultrasound machine [28]. Noticeable are the layered tissue, which produces structured echogenic diversity. Similar to what is seen in the ovine and bovine tissue and dissimilar to what is seen in the phantom. The tumours share similar resemblance to that produced using agar, creating a region of concentrated, higher echogenicity with diffusion along the margins.

**Figure 5.**
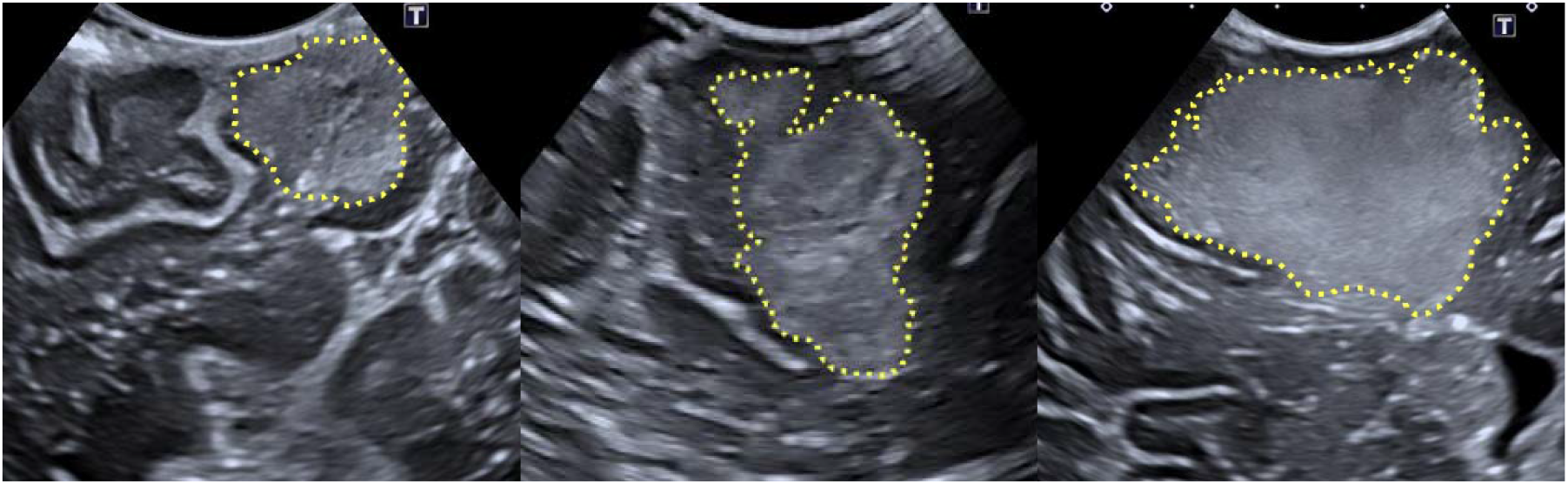
Human intraoperative ultrasound images of Glioblastoma. The tumours are highlighted using the yellow dotted lines. The left and right figures show a maximum depth of 5cm, while the middle figure shows a maximum depth of 4.5cm.

A cross section of the agar in one of the ovine ex vivo samples is shown in Figure.7. The agar was mixed before injection with acriflavine to dye the tissue. As shown in the figure, it is possible to embed the agar solution in an area of normal tissue.

### Necrosis Simulation

Figure.6 shows an example of the outcome of injecting water directly into the agar solution. In this example, the water successfully displaces the agar, creating a nested spherical area of lower echogenicity within the higher echogenic agar solution. This can be seen as an area of slightly higher echogenicity than the background levels. The figure also shows an image of a human brain tumour with a necrotic core to highlight the impact on the echogenic profile.

**Figure 6.**
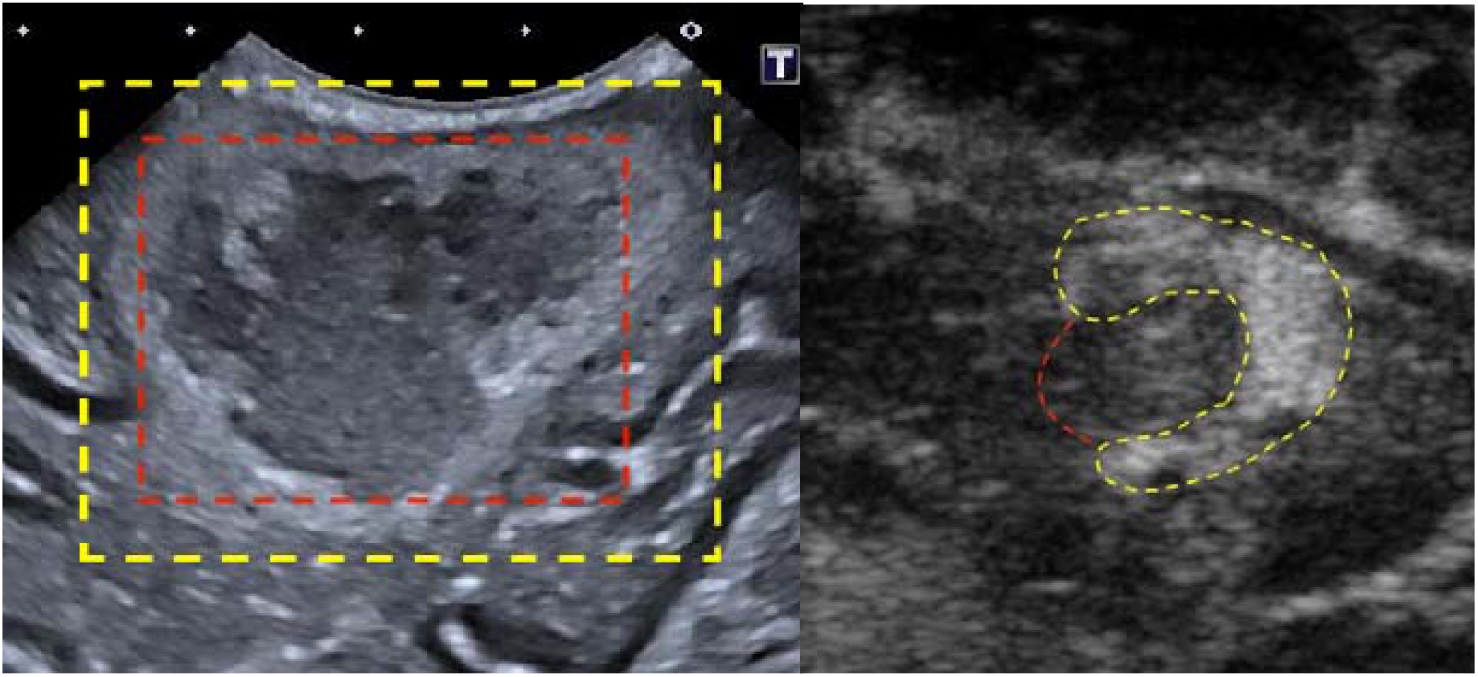
The right example shows a real case of a brain tumour, highlighted in yellow, with a necrotic core, highlighted in red. On the left is water added to the agar to simulate a necrotic core. Yellow line outlines the agar, red line outlines the water.

**Figure 7.**
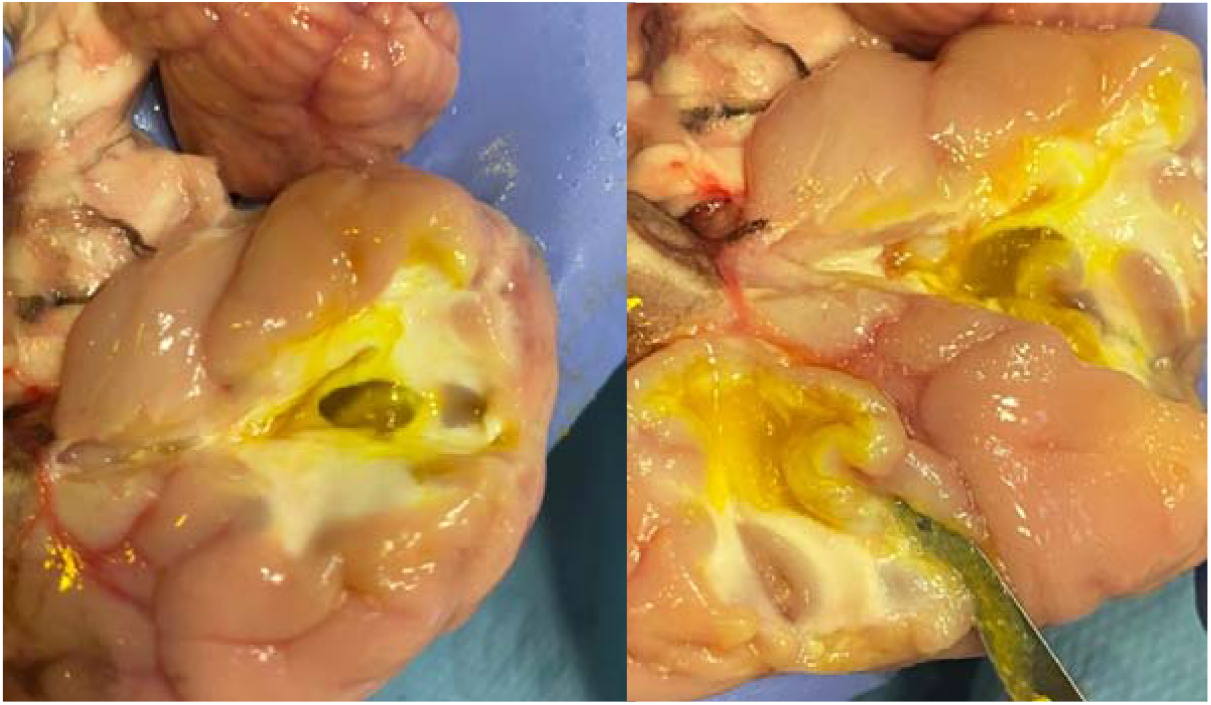
Cross section of one of the ovine brains. In this sample, two sites were injected with agar (dyed yellow)

## Discussion

The creation of resources for surgical simulation, particularly the simulation of anatomy, is ever in demand and requires ongoing commitments to overcome a series of technical, logistical, and regulatory challenges. Regarding the specific problems related to the application of IOUS during brain tumour surgery, the current article addresses one of the critical issues related to this imaging technology, the lack of simulation resources to support preclinical / cadaveric training and technical innovation.

Through our experiments with agar injection into animal ex vivo brain tissue, we show that it is possible to mimic tumours in inexpensive real tissue, replicating an anatomy that is as similar to human as possible and providing the expected ultrasound feedback. The simulation of necrosis, by injecting water into the hardened agar, created an acceptable resemblance to a high-grade real-life tumour scenario. This solution removed a few of the issues related to the artificial phantom, namely the lack of resemblance to the surrounding gyri and sulci, the acoustic shadow underlying the tumour model (absent in most cases) and overall, the signal obtained in terms of both resolution and reliability.

The creation of medical simulation resources such as this is important, as it provides a method of improving practical skills and reducing the dependency on learning from experience during real surgeries. Extending from this, it would also facilitate learning new US-guided techniques such as US-guided core biopsies which are presently, due to lack of access, under applied in neurosurgery but readily used in other medical fields. For example, in this case the operators found that the task of using US to guide the agar injections was qualitatively associated with an improvement in US coordination and US localisation of the needle.

In addition, inexpensive, accurate simulation of brain anatomy and pathology encourages technical innovation. For example, data can be collected more easily (possibly to supplement the more difficult-to-acquire in vivo data), to enable deep learning training, which usually requires large volumes of data to prevent overfitting. Furthermore, the inexpensiveness and anatomical and textural accuracy of the phantom can provide a tool for conducting robotic experiments [16]. As with most robotic applications for surgery, ultrasound guidance is still in an early preclinical, experimental research-focused phase. As such, this has allowed for temporary circumvention of the simulation of more challenging surgical procedures which may utilise ultrasound, and the expected intricacies of the anatomical features that would be present. In a large number of these robotic setups, the application is non-invasive, on-the-skin procedures [17] [18], or are works that can be evaluated on simple synthetic medical phantoms, to explore technical novelty in the design and control of the robotic system [19] [20] [21] [22]. Nevertheless, evaluation of robotic systems on ex vivo is not uncommon. For example human cadavers for spine scanning [23], porcine tissue for needle tracking [24], kidney and heart for learning-from-demonstration [25]. Germane to this article, [26] uses an unmodified chicken breast for the evaluation of focused US ablation. The study pertained to the feasibility of real-time detection of the ablation area and, as such, the task did not require the ablated tissue to be pathological. However, the clinical motivation for this research and many others within the field of robotic ultrasound is therapeutic surgery. By using the method proposed in this paper, or future similar methods, the tumour mimicking tissue can act as the target for the robotic experiment e.g. thermal ablation.

Our solution is not without limitations. First, prion disease represents a serious biohazard and needs to be addressed properly when manipulating the brains of dead animals. We have circumvented this problem opting for young animal brains (veals and lambs) specifically harvested for human consumption and have already gone through official clearance with the relevant regulatory authorities. If an adult animal brain is preferable for whatever reason, a much stricter protocol would be required. As an additional precaution measure to manage any biosafety risk, all biological material has been discarded according to standard safety protocols. Second challenge, working with ex vivo biological tissue should ideally be done in biosafe laboratories, unlike commercial phantoms, which could be used in any setting. This set-up is per se expensive and/or not always accessible for all centres. Finally, while the tissue does show much closer resemblance with the in vivo images, the possibility of degradation-related artefacts must be considered. For these experiments, tissue samples were kept out of the refrigerator at room temperature for more than an hour, and the harvesting time presumably took place 24 to 48 hours earlier. However, no significant necrosis or tissue degradation was observed within this time period, both on macroscopic inspection and on imaging. Some degree of manipulation-related damage was noticed especially at the level of the basal structures, but this was negligible for the purpose of our acquisitions. Some of the tissue that was left out was later frozen and used on another day, and it was observed that the fluctuation of temperature and the freezing process minimally impacted the usability of the tissue. In general, we found that this setting and time frame is acceptable, reliable, and replicable.

## Conclusions

Methods for mimicking tumours within ultrasound images are valuable both for clinical training and research. This is especially the case with brain tumours, which are complex, highly variable, and diffuse. IOUS is a difficult tool to become proficient with, outside of real surgeries. The lack of affordable and accurate commercially available phantoms creates the need for home-made methods to create inexpensive homemade phantoms. Our proposed method solves this by mimicking tumours within real tissue, by combining easy and cheap butchers meat with agar powder. Furthermore, this will encourage technical innovation, as the lack of simulation resources has been a significant bottleneck in technical innovation. A data set is created and made publicly available upon request that contains bounding-box annotations of multiple ex vivo ovine brains containing tumour-mimicking agar. The data set contains 576 annotated images and 388 images without tumours, where the frames were extracted from US videos at 1Hz. The images are stored as MAT-files with the annotations in CSV files with the structure [top left corner, width, height]. Sample images from the data set are shown in Figure.8.

**Figure 8.**
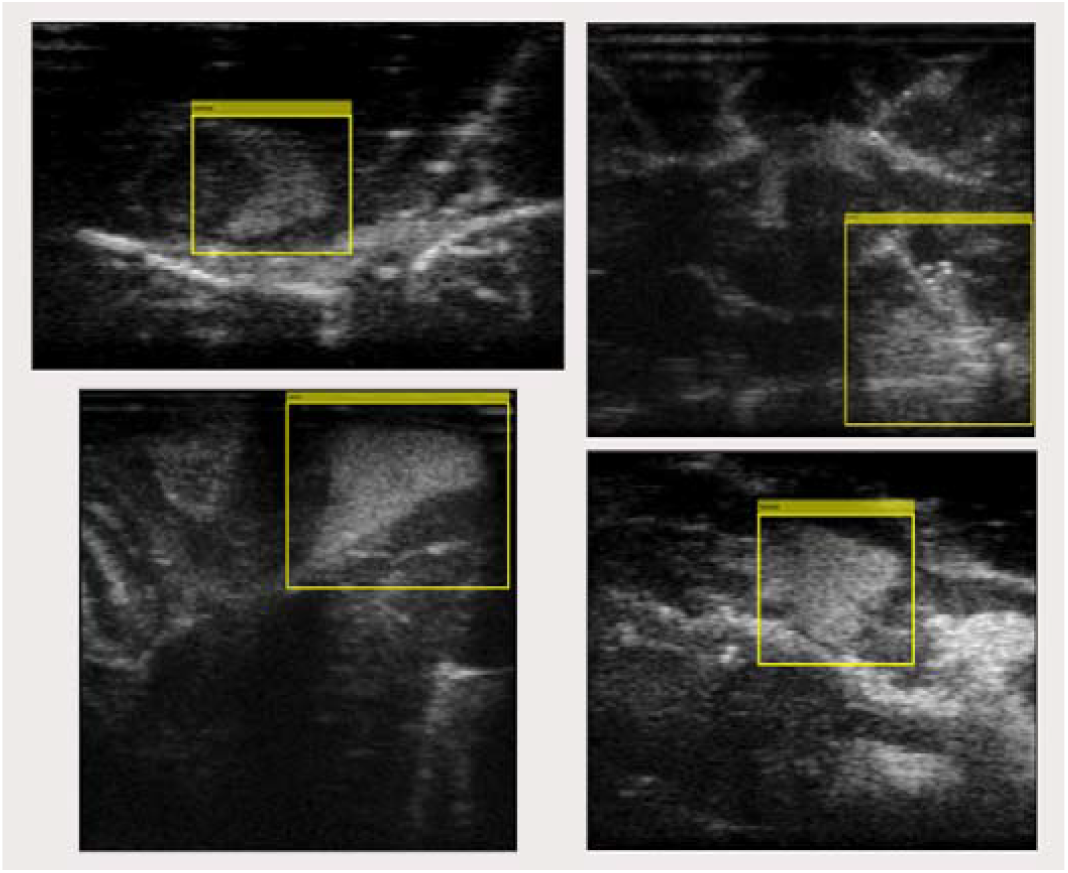
Samples from the data set with super imposed bounding box annotation.

Future work is encouraged to improve the realism of the tumour mimicry. This could include testing different concentrations of agar solution, testing different locations, and using more than one compound to simulate tumour characteristics, creating more complex layering and diffusion effects. Further research is desirable to simulate a more infiltrative type of lesion, with reduced margin sharpness.

## Data Availability

All data produced in the present study are available upon reasonable request to the authors

## Abbreviations

IOUS: Intraoperative ultrasound
US: Ultrasound

## Compliance With Ethical Standards

### Funding

This project was supported by the UK Research and Innovation (UKRI) Centre for Doctoral Training in AI for Healthcare (EP/S023283/1), the Royal Society (URF\R\ 201014), the NIHR Imperial Biomedical Research Centre.

### Ethical Approval

This article does not contain any studies with human participants or animals performed by any of the authors.

